# Multidimensional analysis of preoperative patient-reported outcomes identifies distinct phenotypes in patients booked for total knee arthroplasty: Secondary analysis of the SHARKS registry in a metropolitan hospital department

**DOI:** 10.1101/2023.09.03.23294749

**Authors:** Raquel McGill, Corey Scholes, Stephen Torbey, Lorenzo Calabro

## Abstract

**Background:** Traditional research on total knee arthroplasty (TKA) relies on preoperative patient-reported outcome measures (PROMs) to predict postoperative satisfaction. We aim to identify distinct patient phenotypes among TKA candidates, and investigate their correlations with patient characteristics.

**Methods:** Between 2017-2021, 389 patients with 450 primary knee cases at a metropolitan public hospital were enrolled in a clinical quality registry. Demographics, clinical data, and the Veterans Rand 12 (VR-12) and Oxford Knee Score (OKS) were collected. Imputed data were utilised for the primary analysis, employing k-means clustering to identify four phenotypes. ANOVA assessed differences in scores between clusters, and nominal logistic regression determined relationships between phenotypes and patient age, sex, body mass index, and laterality.

**Results:** The sample comprised 389 patients with 450 primary knees. Phenotype 4 (Mild symptoms with good mental health) exhibited superior physical function and overall health. In contrast, patients in phenotype 2 (Severe symptoms with poor mental health) experienced the most knee pain and health issues. Phenotype 1 (Moderate symptoms with good mental health) reported high mental health scores despite knee pain and physical impairment. Patient characteristics significantly correlated with phenotypes; those in the Severe symptoms with poor mental health phenotype were more likely to be younger, female, have a higher BMI, and bilateral osteoarthritis (P<0.05).

**Conclusions:** This multidimensional analysis identified TKA patient phenotypes based on common PROMs, revealing associations with patient demographics. This approach has the potential to inform prognostic models, enhancing clinical decision-making and patient outcomes in joint replacement.

**Significance and Innovations:** This study leverages the power of machine learning to simultaneously analyse multiple patient-reported outcome measures, which is not utilised in traditional research in total knee arthroplasty

Four distinct phenotypes were identified, and they demonstrated significant associations with patient demographics

This method has potential for developing prognostic models in joint replacement, with the ability to improve clinical decision-making and patient outcomes.

## Introduction

Total knee arthroplasty (TKA) has been instrumental in improving the quality of life for patients with advanced osteoarthritis (OA) (1). Globally, OA affected 595 million people in 2020 and is projected to rise by 74.9% for knee OA by 2050 (2). Anticipated primary TKA demand is projected to rise by 276% by 2030 in Australia, with economic implications estimated at $1.38 billion (3). Comparable rises in arthroplasty have been projected in the USA, UK, Canada, Denmark, Sweden, New Zealand (4–9). Despite the overall success of TKA, on average10% of patients report dissatisfaction after surgery (10,11). The multifactorial reasons behind this dissatisfaction are not fully understood, presenting a challenge for even experienced surgeons to accurately predict clinically meaningful improvements after joint replacement (12,13). Gaining a greater understanding of preoperative patient factors would enable surgeons to better select, and more effectively counsel patients prior to surgery.

Previous research has shown that patient-reported outcome measures (PROMs) and demographic data influence satisfaction rates (14). Variables such as preoperative levels of pain and function, age, sex, body mass index (BMI), mental health, opioid use, and comorbidities have shown varying degrees of association with outcomes (14–16). Notably, preoperative PROMs assessing knee-specific parameters and mental health have the strongest association with postoperative outcomes (14). However, these findings and approaches have yet to achieve clinically significant reliability (17).

Knee OA’s marked heterogeneity in pathology, progression, demographics, and treatment response likely complicates outcome prediction, prompting consideration of whether patient variability can be explained by discrete subgroups or phenotypes (18,19). Phenotypes refer to groups of patients with similar characteristics, such as demographics, clinical presentation, disease progression, and treatment response. Recent studies have found that phenotypes based on a combination of preoperative PROMs were associated with dissatisfaction after TKA and postoperative PROMs (20,21). These studies highlight the utility of combining patient data to create a holistic view of patients compared to the traditional evaluation of multiple PROM independently.

An emerging approach to identify phenotypes based on a combination of patient data involves the use of machine learning. Cluster analysis is an unsupervised machine learning technique that can simultaneously analyse and combine scores from various PROMs to identify more nuanced patterns. Kung et al. (21) used cluster analysis to identify phenotypes based on preoperative PROMs and found associations with TKA outcomes. They used a PROM called Patient Reported Outcome Instrumentations System, which measures physical, mental, and social well–being in the general patient population. While this is a validated tool, it is not specific to orthopaedic research and is not a routinely collected PROM by clinicians.

To the best of our knowledge, previous research has yet to leverage cluster analysis to identify phenotypes in patients undergoing TKA using knee-specific and general well-being PROMs. Therefore, this paper aims to i) establish the feasibility of identifying phenotypes in patients with OA booked for TKA based on simultaneous cluster analysis of multiple preoperative PROMs and ii) evaluate the association between these phenotypes and patient demographics. It was hypothesised that distinct phenotypes would be identifiable within the multidimensional PROMs data, and associated with specific patient demographic factors.

## Patients and Methods

### Study Design

Secondary analysis of a prospectively collected, clinical registry within a hospital department. The study was reported in line with RECORD guidelines (Supplementary A) <u>(Benchimol et al. 2011)</u>.

### Setting

The data was extracted from a registry within a single hospital, which is the main centre for elective joint replacement within the local health district of a state capital city within Australia.

### Patient selection

Patients booked for primary TKA between 2017 and 2023 at a metropolitan public hospital were enrolled in the Shoulder, Hip, Arthroplasty and Knee Surgery (SHARKS) clinical quality registry. This is a registered (ANZCTRN: 12617001161314), clinical quality registry with Human Research Ethics approval (HREC/16/QPAH/732) administered by the Department of Orthopaedics, with contributions from the Department of Physiotherapy. Patients who provided consent to be part of the registry were enrolled in the study.

### Variables

The outcome of this study analysis was the phenotypes identified within the patient sample using routinely collected patient-reported outcome measures. Demographic predictors were identified as age at initial examination, sex, body mass index and bilateral status (presenting with one or both knees for surgery). Quantitative variables were treated as continuous in all analyses without dichotomisation into categories.

### Data Sources

The registry collects demographic data (including postcode) and clinical data, as well as patient-reported outcomes before and after surgery (22). Clinical data for this analysis included the type of surgery (primary or revision) for case selection and bilateral status. The two PROMs used for this study were the Veterans Rand 12 (VR-12) and Oxford Knee Score (OKS). These PROMs were selected due to their efficiency in capturing multiple domains of general health (VR-12) and knee-specific pain and function (OKS). Alternative PROMs such as the Knee Injury and Osteoarthritis Outcome Score (KOOS) and Western Ontario and McMaster Universities Osteoarthritis Index were rejected due to their patient burden in the specific hospital setting. The VR-12 is a 12-item questionnaire that measures health-related quality of life and estimates disease burden (23). The VR-12 was subdivided into the physical (VR-12 PCS) and mental (VR-12 MCS) component scores. The OKS is a 12-item questionnaire specifically designed and developed to assess function and pain within the past 4 weeks following a TKA, but has also been shown to be a reliable and valid tool for measurement of symptoms in patients with OA (24). The OKS was subdivided into its functional (OKS-F) and pain (OKS-P) subscales, expressed on a scale from 0 to 100 (worst to best) (24). Preoperative data was collected on paper forms at the time of booking for surgery in the clinic.

### Bias and Study Size

The sample was one of convenience extracted from the department registry for total knee replacement at the time of analysis. Clustering in itself is not directly impacted by sample size with respect to power (25). Missing data was addressed with a multiple imputation approach to avoid the inherent bias that can occur with complete case analysis.

### Data Access and Cleaning

The database was wholly accessible to the authors for the duration of the study. The SHARKS registry undergoes routine monitoring and cleaning as previously described (22). Data was retrieved from a routine export from the registry that is produced every three months. The export used in this analysis included all cases captured into the registry up to 31st of October 2023. The export was accessed in the R environment using RStudio (v2023.12.0, Build 369) and all cleaning, processing and analysis performed using dedicated packages. The code and explanatory notes, as well as relevant citations were compiled in an analysis report (Supplementary B).

### Missing Data

Missing data for BMI, OKS-P, OKS-F, VR12-MCS, and VR12-PCS were imputed in order with sequential chained equations based on partial mean matching drawing on a pool of 3 nearest neighbours (k) for a total of 20 (m) imputed datasets. Posterior estimates of model parameters were obtained using sampling replacement (bootstrap). The variables included in the imputation model were:

● Age at surgery (years)
● Sex (male; female)
● Bilateral status (bilateral; unilateral)
● Interval of surgery date from first case in the sample (days)
● Estimated median annual income based on postcode ($/year). Retrieved from the Australian Tax Office (26)

The imputation process was performed using the *mice package* (Supplementary B pg 6)) under the assumption that the data contained missingness at random considering the collection methods in place from the inception of the registry. The imputed data was used for the primary analysis. A summary of the missing data can be found in Supplementary B.

### Data and Statistical Analysis

Sample descriptives and demographics were presented as means and standard deviations for continuous variables, while percentages were provided for categorical variables. Patients responding similarly to questionnaires were assessed using clustering across multiple subscales. The VR-12 MCS, VR-12 PCS, OKS-F, and OKS-P subscale scores were used. A k-means clustering algorithm was applied to the PROMs subscales (Supplementary B pg 8). This iterative process partitions data into groups based on the distance to group centroids (distance to a cluster mean). The chosen value k determines the number of phenotypes the algorithm will form. A dedicated package (miclust) was used to apply the elbow method for optimal k selection using between-cluster sum of squares as the indicator. This approach identified k = 3 as the optimal k. For this study, a k value of 4 was chosen to test the cluster concept and to take advantage of a slight reduction in within-cluster variability. . Due to the imputed dataset, the phenotype in which each case was allocated was determined by extracting the mode average across the 20 imputations for each individual. Regression analyses were conducted by combining data from the multiple imputed datasets, to compare the PROM subscales between phenotypes. Multinomial logistic regression was performed to assess the relationship between phenotype and patient demographics (age, BMI, sex) and bilateral status. The reference category was selected based on review of the patterns in the PROMs subscales across phenotypes. Odds ratios were calculated to enable ease of interpretation. Alpha was set a-prior at 5% to identify significant results.

## Results

### Patient characteristics and overall PROMs

There were 450 cases assessed for eligibility, and after exclusions for missing data (examination date and revision procedures), there were 377 patients and 432 primary knees included in the study. Patient demographics and preoperative PROMs for the cohort are summarised in Table 1. The patient cohort were on average 69 years old and classified as obese class I with 53.6% females. There were 33.2% of patients booked for bilateral TKA (95% CI: 28.9 - 37.8). In this cohort, primary osteoarthritis accounted for 96% of TKA cases, while the remaining comprised rheumatoid arthritis and post-traumatic arthritis.

**Table 1.**
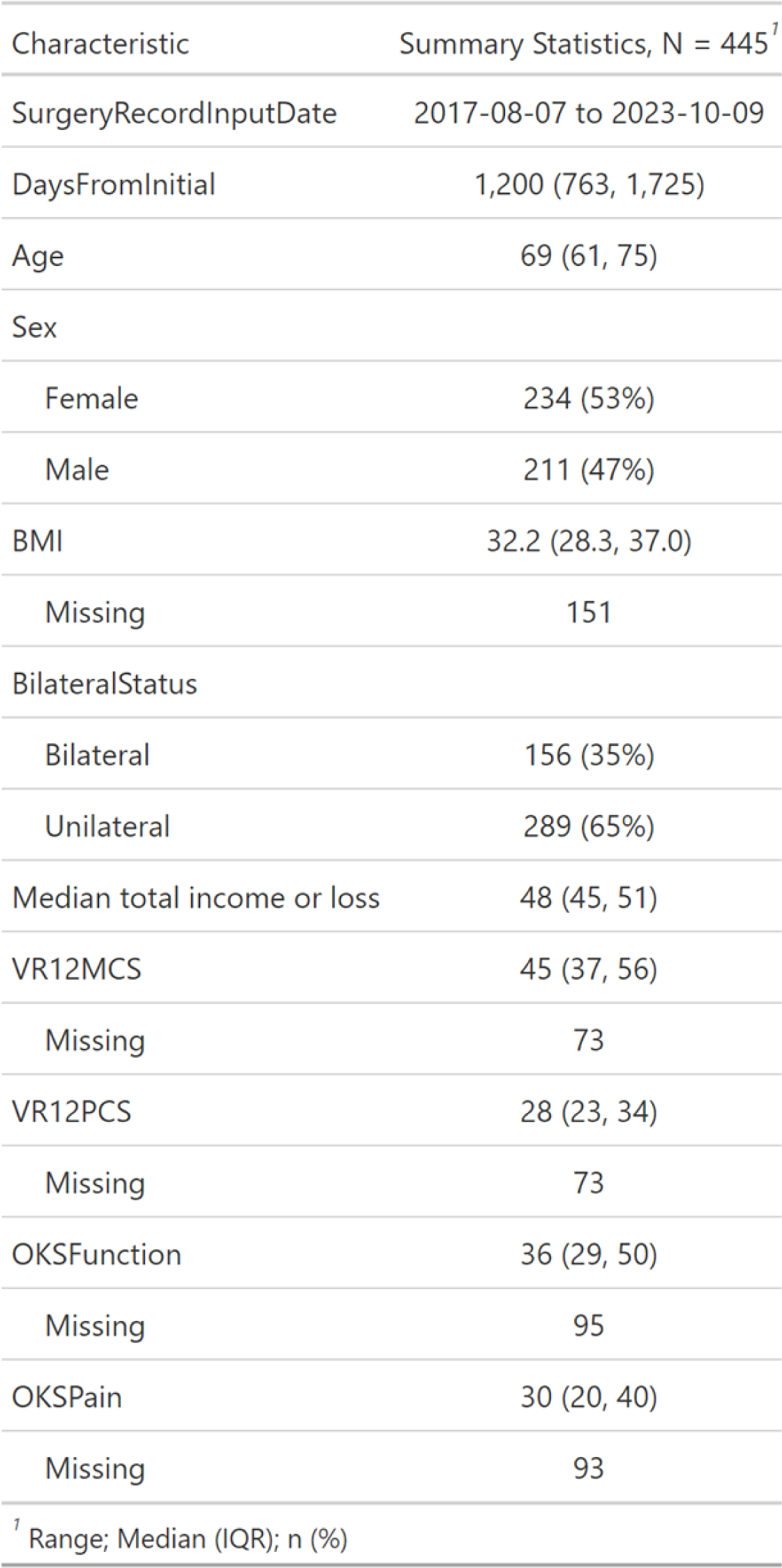
Patient demographics and preoperative PROMs

### Phenotypes

The model partitioned patients into one of four phenotypes based on their preoperative PROMs scores. The phenotype labelling remained stable in the majority of the sample across imputations (Supplementary B pg13). The degree of functional impairment, pain severity, and general well-being reported by the patients in the present cohort existed along a spectrum . However, the patients belonged to discrete phenotypes based on these preoperative scores. The differentiation among phenotypes is visualised in a three-dimensional scatter plot, which uses OKS-P, OKS-F, and VR12-MCS (Figure 1).

**Figure 1.**
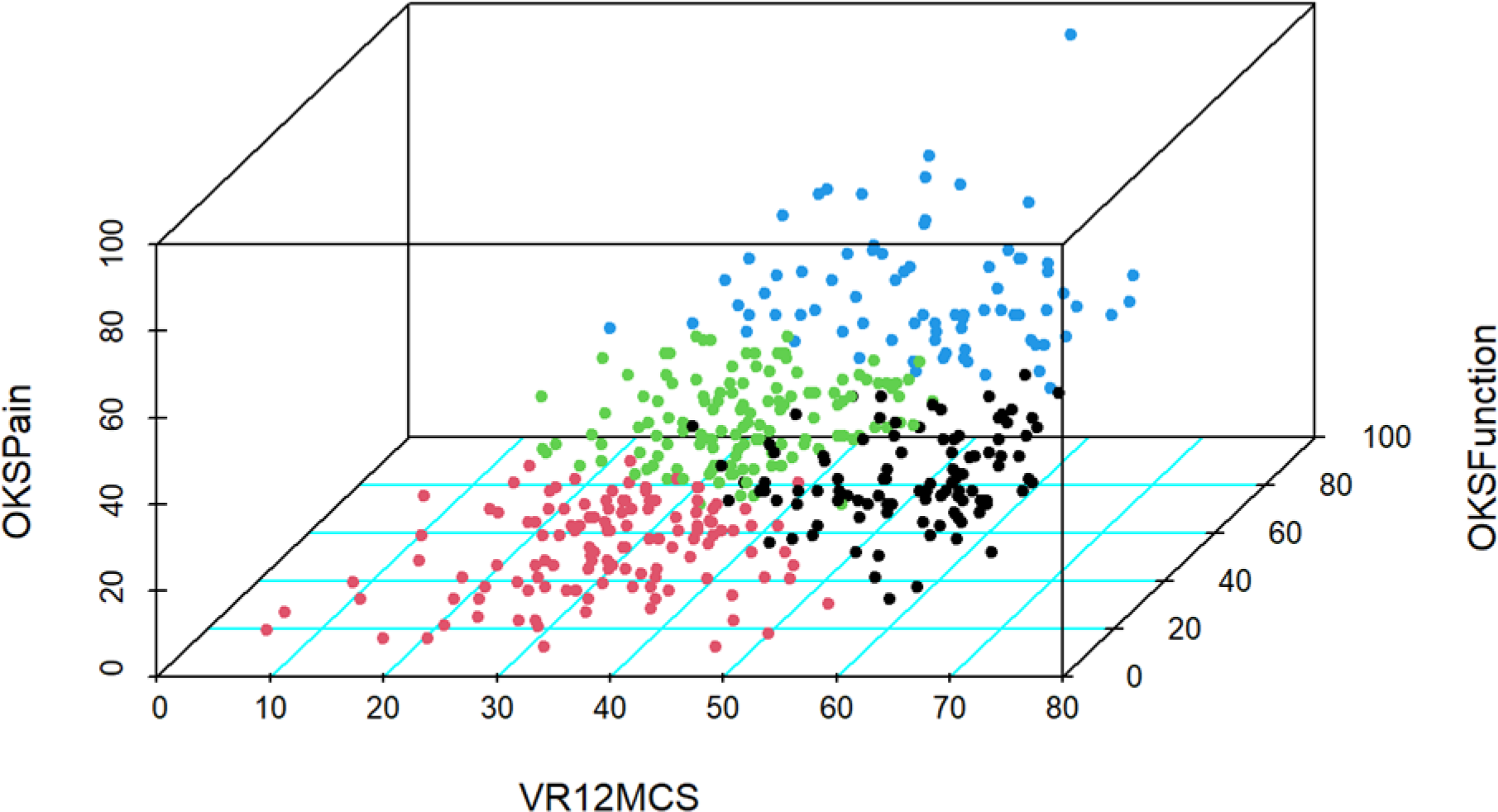
A 3D scatter plot demonstrating each patient’s scores in the OKS-F, OKS-P, and VR-12 MCS. Patients were classified into their phenotype which is demonstrated based on the following colours: phenotype 1 = black ; phenotype 2 = red; phenotype 3 = green; and phenotype 4 = blue. VR12-PCS data not shown.

The four phenotypes represent distinguishable patterns of responses to each of the four PROMs (Figure 2). Figure 2 demonstrates the adjusted mean values for each PROMs subscale for the four phenotypes. Phenotype 4 scored the overall highest across the four PROMs subscales, while phenotype 2 scored the overall lowest. Interestingly, phenotype 1 scored the second worst overall on the OKS-P, OKS-F, and VR-12 PCS subscales, while scoring the highest on the mental well-being component on the VR-12 compared to all other groups. Phenotype 3 scored average on the PROMs subscales compared to the other four phenotypes. These four phenotypes can be characterised as the following (Figure 3): *Moderate symptoms with fair mental health* (phenotype 3, N = 132), *Severe symptoms with poor mental health* (phenotype 2, N = 80), Mild symptoms with good mental health (phenotype 4, N = 106), and Moderate symptoms with good mental health (phenotype 1, N = 127).

**Figure 2.**
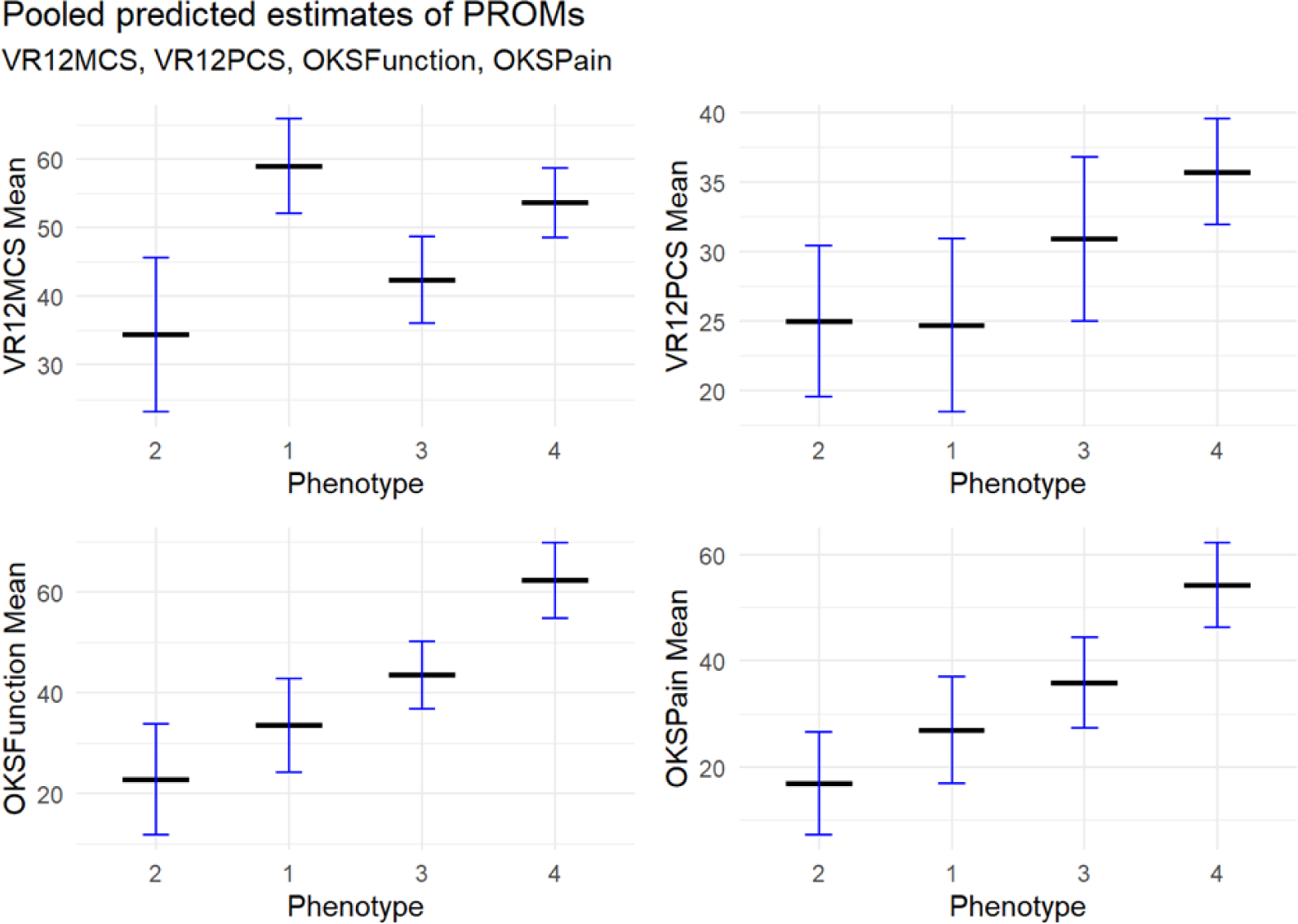
Model-predicted means (with 95% confidence intervals) for each PROM across phenotypes.

**Figure 3.**
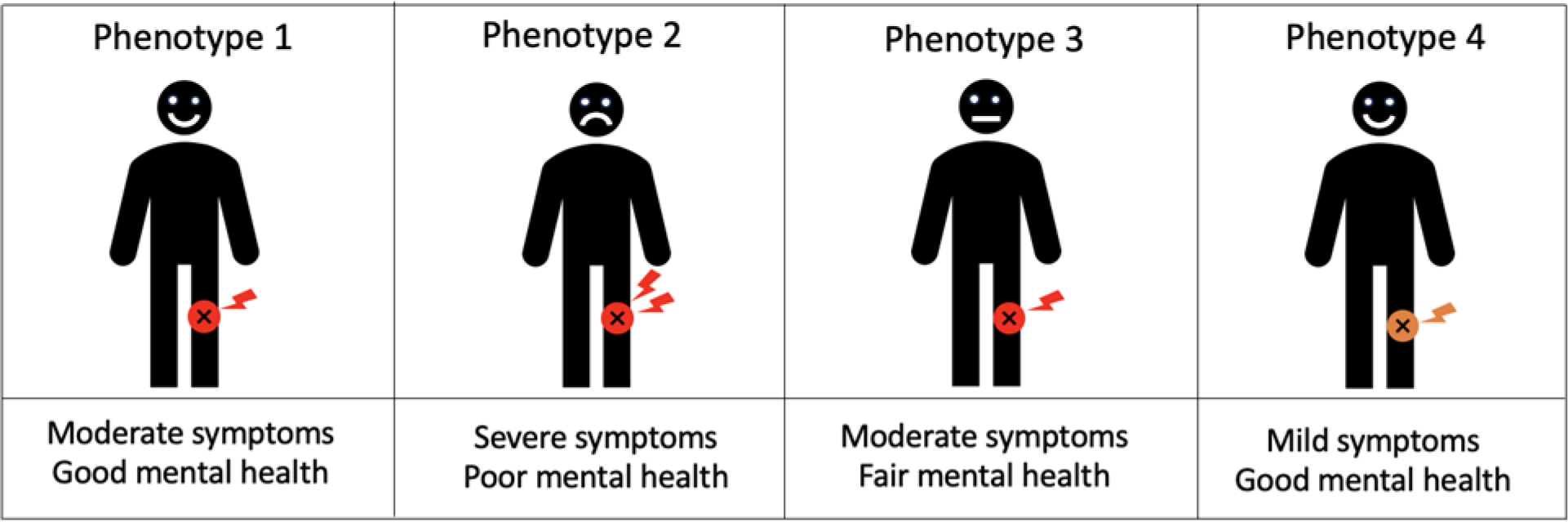
Graphic representation of the four phenotypes identified by cluster analysis. Moderate symptoms with good mental health (phenotype 1), Severe symptoms with poor mental health (phenotype 2), Moderate symptoms with fair mental health (phenotype 3), Mild symptoms with good mental health (phenotype 4)

### Demographic Relationships to Phenotype

Age, BMI, sex, and laterality demonstrated significant (P<0.05) relationships with phenotypes (Table 2). Those in the *Severe symptoms with poor mental health* phenotype (phenotype 2) were more likely to be younger patients with a greater BMI (Table 3). These patients were also more likely to be female and have bilateral presentations of knee OA (Table 2, Table 3). Conversely, patients in the *Mild symptoms with good mental health* phenotype (phenotype 4) were more likely to present with a lower BMI compared to the *Moderate symptoms with good mental health* phenotype (phenotype 3). Additionally, the phenotypes reporting good mental health were more likely to be older.

**Table 2.**
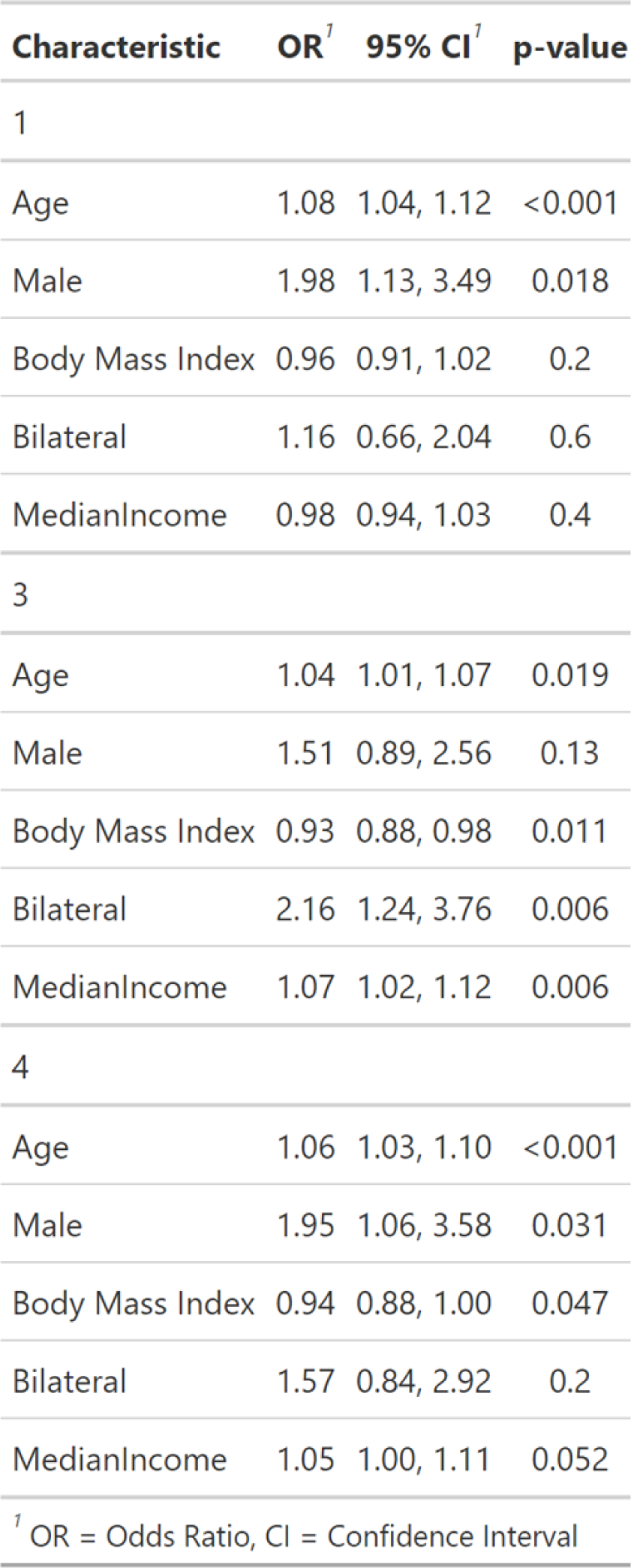
Summary of patient demographics with phenotype membership (compared to phenotype 2)

**Table 3.**
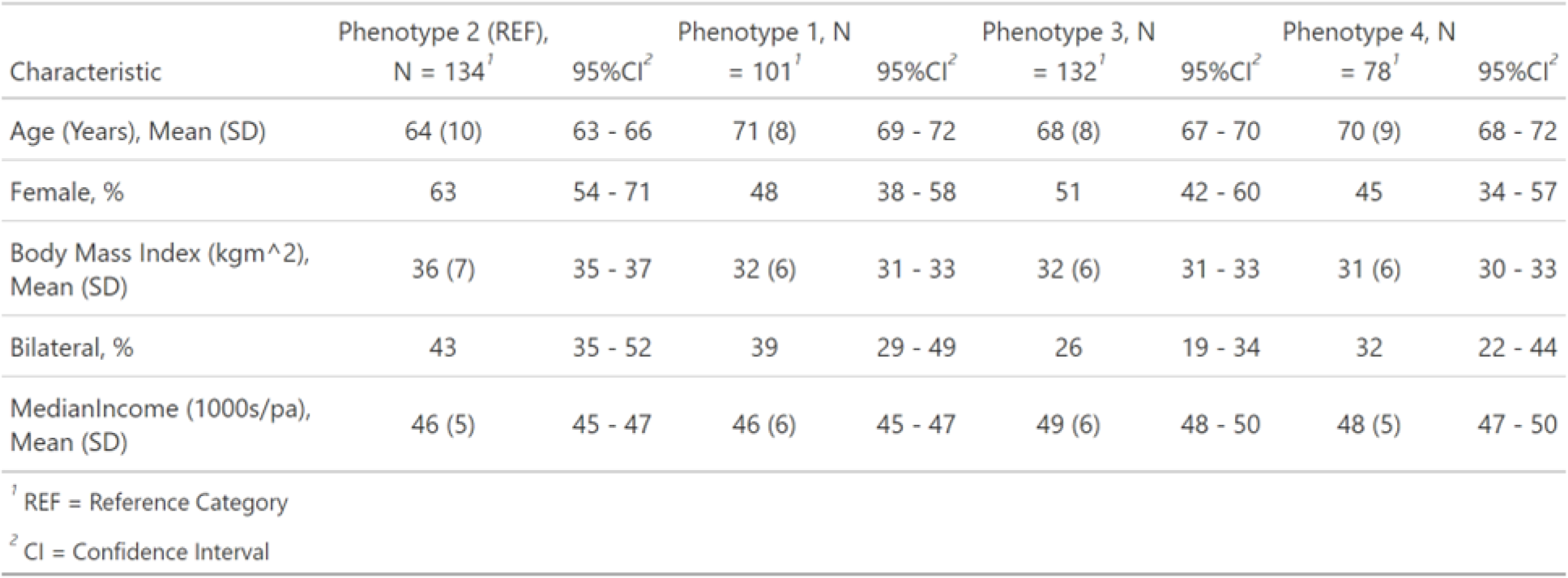
Predicted means of BMI and Age and proportions of Bilateral status and Sex (with 95% confidence intervals) for each phenotype.

| Characteristic | Phenotype 2 (REF), |  | Phenotype 1, N |  | Phenotype 3, N |  | Phenotype 4, N |  |
| --- | --- | --- | --- | --- | --- | --- | --- | --- |
|  | N = 134<sup>1</sup> | 95%CI<sup>2</sup> | = 101<sup>1</sup> | 95%CI<sup>2</sup> | = 132<sup>1</sup> | 95%CI<sup>2</sup> | = 78<sup>1</sup> | 95%CI<sup>2</sup> |
| Age (Years), Mean (SD) | 64 (10) | 63 - 66 | 71 (8) | 69 - 72 | 68 (8) | 67 - 70 | 70 (9) | 68 - 72 |
| Female, % | 63 | 54 - 71 | 48 | 38 - 58 | 51 | 42 - 60 | 45 | 34 - 57 |
| Body Mass Index (kgm<sup>2</sup>), Mean (SD) | 36 (7) | 35 - 37 | 32 (6) | 31 - 33 | 32 (6) | 31 - 33 | 31 (6) | 30 - 33 |
| Bilateral, % | 43 | 35 - 52 | 39 | 29 - 49 | 26 | 19 - 34 | 32 | 22 - 44 |
| MedianIncome (1000s/pa), Mean (SD) | 46 (5) | 45 - 47 | 46 (6) | 45 - 47 | 49 (6) | 48 - 50 | 48 (5) | 47 - 50 |

<sup>1</sup> REF = Reference Category
<sup>2</sup> CI = Confidence Interval

## Discussion

This study used a multidimensional cluster analysis approach to identify phenotypes in preoperative TKA patients and to evaluate the association between these phenotypes and demographics. Our findings indicate that patients with end-stage knee OA form a heterogeneous group, exhibiting varying degrees of functional impairment, pain severity, mental health, and general physical well-being. Simultaneous analysis of OKS-F, OKS-P, VR-12 MCS, and VR-12 PCS scores revealed patterns in preoperative PROMs that suggest patients in our cohort belong to unique phenotypes.

The cluster analysis successfully identified four distinct phenotypes that demonstrated clinical correlations with demographic factors. These subgroups were categorised as *Moderate symptoms with good mental health* (phenotype 1), *Mild symptoms with good mental health* (phenotype 4), *Moderate symptoms with fair mental health* (phenotype 3), and *Severe symptoms with poor mental health* (phenotype 2). Interestingly, these four phenotypes are similar to a previous study that used a combination of preoperative PROMs to identify subgroups in patients undergoing TKA. The four phenotypes they identified were *Normal function*, *Mild impairments*, *Impaired with distress*, and *Impaired without distress* (21). The similarity between the phenotypes supports the concept that there exists discrete subgroups of patients with a spectrum of physical symptoms and mental well-being.

The *Mild symptoms with good mental health* phenotype (phenotype 4) had the highest scores for the VR-12 PCS, OKS-F, and OKS-P subscales, and second highest for the VR-12 MCS subscale. These patients rated themselves as having high levels of function, less pain, and better general physical and mental health compared to the majority of patients. In contrast, patients in the *Severe symptoms with poor mental health* phenotype (phenotype 2) reported the lowest scores for the VR-12 PCS, VR-12 MCS, OKS-F, and OKS-P subscales, indicating worse knee pain, impairment, and general physical and mental health in comparison to the other patients. In the present cohort, patients who reported poor mental health also reported the lowest knee-specific and general health PROMs scores. However, not all patients experiencing moderate knee pain and functional impairment reported compromised mental health. This highlights the complex interplay between mental health and a patient’s perspective on their knee pain. A unique combination of PROMs scores was observed in the *Moderate symptoms with good mental health* phenotype (phenotype 1), where patients had the second lowest scores in the VR-12 PCS, OKS-P, and OKS-F subscales but the highest VR-12 MCS scores compared to all other phenotypes. These patients reported the highest mental health scores despite having significant knee pain, functional impairment, and poor general physical health, The notable disparity in self-reported mental well-being between the phenotype 2 and phenotype 1 raises interesting speculations regarding their postoperative outcomes.

Indeed, a previous study reported phenotypes with high levels of mental well-being yet significant knee pain and impairment, were associated with better postoperative satisfaction compared to phenotypes with both poor mental health and significant knee pain and impairment (20). This underscores the importance of adopting a holistic approach to patient care and considering factors such as preoperative depression, catastrophising tendencies, and anxiety, as they are associated with TKA dissatisfaction (11,27). Therefore, patients eligible for TKA identified with poor mental health may benefit from additional support such as educational programs, cognitive–behavioural therapy, and exercise therapy with graded activity levels (28).

Further analysis revealed that the demographics associated with phenotypes were aligned with the current literature. Patients who were female, younger, and had a higher BMI were more likely to be in the *Severe symptoms with poor mental health* phenotype (phenotype 2), which had the lowest scores for the VR-12 PCS, VR-12 MCS, OKS-F, and OKS-P subscales. Moreover, patients in this group were more likely to have both knees booked for TKA. The demographic factors that were associated with worse preoperative PROMs in this cohort have been shown to correlate with poor postoperative outcomes. The multidimensional analysis employed in this study provides a valuable alternative for identifying phenotypes that have significant clinical correlations supported by literature.

Phenotyping is an emerging field in orthopaedic research that offers a more nuanced understanding of patient heterogeneity and has the potential to guide management decisions (29). While this study focused on PROMs, previous research has explored OA phenotypes has attempted to group patients with OA into phenotypes based on gait kinematics (20,21), joint aspirate metabolomics (22), pain groups (23), functional knee characteristics (24), structural characteristics, and prognosis (25). For instance, Moser et al. (2020) identified diverse knee phenotypes based on knee alignment and recommended personalised TKA realignment for improved outcomes. By identifying distinct patient phenotypes, researchers can investigate the underlying variables that contribute to disease progression, prognosis, and outcomes (18). While the prognostic value of the phenotypes identified in this study are yet to be determined, these phenotypes may aid in refining patient selection, targeted preoperative interventions, and enhancing shared decision-making with patients presenting for surgical review of OA.

While phenotyping has the potential to offer significant benefits to orthopaedic research and clinical practice, caution is necessary in regards to the phenotypes themselves and the analytical approaches used to develop them (18). A number of different phenotypes could be identified based on the variables included in the model irrespective of their true relevance for phenotyping. A systematic review suggested that omitting patient data from different domains, such as clinical, imaging, or laboratory data, may hamper the conclusions drawn from phenotypes (30). To further complicate the endeavour, adding an increasing number of variables to the model may not necessarily equate to greater accuracy (30). Overall, while phenotypes offer an exciting alternative approach, caution is necessary to improve the accuracy and applicability of these approaches to orthopaedic research and clinical practice.

### Limitations

The generalisability of the findings in this study is impacted by the patient characteristics within the cohort. It is challenging to determine whether the majority of patients with OA booked for TKA would fit into one of the identified phenotypes. Future research is necessary to assess the stability of the identified phenotypes, validate the results in different independent cohorts, and examine longitudinal outcomes. Although multiple imputation was used to estimate missing data and mitigate potential bias, it may have influenced the cluster analysis and generalisability. Finally, the composite nature of the PROMs used in this study poses challenges in their clinical application, as combining scores into nominal variables for labelling groups may introduce accuracy issues and information loss (31). Clinical application of PROMs data is challenging, but a model-based approach may offer avenues to effectively use this information to support shared-decision making and resource allocation, particularly in public health settings where optimisation is crucial.

## Conclusion

This preliminary study used a multidimensional cluster analysis approach to identify phenotypes in end-stage knee OA patients based on commonly used PROMs. These phenotypes were significantly associated with patient demographics in a clinically resonant way. An advantage of this approach is the ability to identify phenotypes that may not be discernible by interpreting PROMs separately. While the predictive utility of these phenotypes is yet to be determined, the use of multidimensional data analysis offers an alternative approach to prognostic models for joint replacement and represents a promising area for further research. The present study provides a foundation for future investigations aimed at refining and validating these phenotypes for the potential use in clinical decision-making and improving patient outcomes.

## Declarations

### Ethical approval and consent to participate

Ethical approval for the registry was obtained from the Human Research Ethics Committee of Metro South Health (Reference HREC/16/QPAH/732), and the registry is registered with the Australian New Zealand Clinical Trials Registry (ACTRN12617001161314).

## Authors contributions

CS was responsible for the study design, statistical analysis, and interpretation of data for the study. RM prepared the manuscript draft and assisted with the interpretation of results. ST performed the original interpretation of analysis and assisted with data collection and interpretation of results. LC was responsible for clinical data collection, providing stewardship to the study, and supervising the manuscript writing. All authors were involved in providing critical review of the manuscript with technical or clinical input and agreement for final approval of the version to be published.

## Data Availability

All data produced in the present study are available upon reasonable request to the authors

## Acknowledgements

The authors thank Andrew Klissanin and Ken Morrissey at QEII Jubilee Hospital, Brisbane, as well as Richard Near and Milad Ebrahimi at EBM Analytics, Sydney for their assistance with this study. The authors also gratefully acknowledge all contributors to the SHARKS registry, including the staff and patients.

