## Supplementary A for "Multidimensional analysis of preoperative patient-reported outcomes identifies distinct phenotypes in patients booked for total knee arthroplasty: Secondary analysis of the SHARKS registry in a metropolitan hospital department"

**The RECORD statement – checklist of items, extended from the STROBE statement, that should be reported in observational studies using routinely collected health data.**

| | Item No. | STROBE items | Location in manuscript where items are reported | RECORD items | Location in manuscript where items are reported |
|---|---|---|---|---|---|
| **Title and abstract** | | | | | |
| | 1 | (a) Indicate the study's design with a commonly used term in the title or the abstract (b) Provide in the abstract an informative and balanced summary of what was done and what was found | | RECORD 1.1: The type of data used should be specified in the title or abstract. When possible, the name of the databases used should be included.<br><br>RECORD 1.2: If applicable, the geographic region and timeframe within which the study took place should be reported in the title or abstract.<br><br>RECORD 1.3: If linkage between databases was conducted for the study, this should be clearly stated in the title or abstract. | Database name included in abstract<br><br>Geography and timeframe outline in abstract<br><br>Not applicable - no data linkage performed |
| **Introduction** | | | | | |
| Background rationale | 2 | Explain the scientific background and rationale for the investigation being reported | | | Included throughout introduction |
| Objectives | 3 | State specific objectives, including any | | | Objectives and hypotheses |

| | | | | | |
|---|---|---|---|---|---|
| | | prespecified hypotheses | | | stated at the bottom of introduction |
| **Methods** | | | | | |
| Study Design | 4 | Present key elements of study design early in the paper | | | Included under relevant heading |
| Setting | 5 | Describe the setting, locations, and relevant dates, including periods of recruitment, exposure, follow-up, and data collection | | | Included under "Setting" and "Patient selection" headers |

| | | | | | |
|---|---|---|---|---|---|
| Participants | 6 | *(a) Cohort study*- Give the eligibility criteria, and the sources and methods of selection of participants. Describe methods of follow-up<br>*Case-control study*- Give the eligibility criteria, and the sources and methods of case ascertainment and control selection. Give the rationale for the choice of cases and controls<br>*Cross-sectional study*- Give the eligibility criteria, and the sources and methods of selection of participants<br><br>*(b) Cohort study*- For matched studies, give matching criteria and number of exposed and unexposed<br>*Case-control study*- For matched studies, give matching criteria and the number of controls per case | | RECORD 6.1: The methods of study population selection (such as codes or algorithms used to identify subjects) should be listed in detail. If this is not possible, an explanation should be provided.<br><br>RECORD 6.2: Any validation studies of the codes or algorithms used to select the population should be referenced. If validation was conducted for this study and not published elsewhere, detailed methods and results should be provided.<br><br>RECORD 6.3: If the study involved linkage of databases, consider use of a flow diagram or other graphical display to demonstrate the data linkage process, including the number of individuals with linked data at each stage. | Study population selection outlined under "Patient selection"<br><br>Coding not used for patient selection in this analysis<br><br>No database linkage used in this study |

| | | | | | |
|---|---|---|---|---|---|
| Variables | 7 | Clearly define all outcomes, exposures, predictors, potential confounders, and effect modifiers. Give diagnostic criteria, if applicable. | | RECORD 7.1: A complete list of codes and algorithms used to classify exposures, outcomes, confounders, and effect modifiers should be provided. If these cannot be reported, an explanation should be provided. | Described under matching header |
| Data sources/ measurement | 8 | For each variable of interest, give sources of data and details of methods of assessment (measurement). Describe comparability of assessment methods if there is more than one group | | | Described under matching header<br><br>Not applicable - only one group examined |

| | | | | | |
|---|---|---|---|---|---|
| Bias | 9 | Describe any efforts to address potential sources of bias | | | Described under matching header |
| Study size | 10 | Explain how the study size was arrived at | | | Described under matching header |
| Quantitative variables | 11 | Explain how quantitative variables were handled in the analyses. If applicable, describe which groupings were chosen, and why | | | Treatment described under “Variables” header |

| | | | | | |
|---|---|---|---|---|---|
| Statistical methods | 12 | (a) Describe all statistical methods, including those used to control for confounding<br>(b) Describe any methods used to examine subgroups and interactions<br>(c) Explain how missing data were addressed<br>(d) *Cohort study*- If applicable, explain how loss to follow-up was addressed<br>*Case-control study*- If applicable, explain how matching of cases and controls was addressed<br>*Cross-sectional study*- If applicable, describe analytical methods taking account of sampling strategy<br>(e) Describe any sensitivity analyses | | | |
| Data access and cleaning methods | | .. | | RECORD 12.1: Authors should describe the extent to which the investigators had access to the database population used to create the study population. | Described under matching heading |

| | | | | | |
|---|---|---|---|---|---|
| | | | | RECORD 12.2: Authors should provide information on the data cleaning methods used in the study. | |
| Linkage | | .. | | RECORD 12.3: State whether the study included person-level, institutional-level, or other data linkage across two or more databases. The methods of linkage and methods of linkage quality evaluation should be provided. | Not applicable - no linkage performed |
| **Results** | | | | | |
| Participants | 13 | (a) Report the numbers of individuals at each stage of the study (*e.g.*, numbers potentially eligible, examined for eligibility, confirmed eligible, included in the study, completing follow-up, and analysed)<br>(b) Give reasons for non-participation at each stage.<br>(c) Consider use of a flow diagram | | RECORD 13.1: Describe in detail the selection of the persons included in the study (*i.e.,* study population selection) including filtering based on data quality, data availability and linkage. The selection of included persons can be described in the text and/or by means of the study flow diagram. | Described under “Patient characteristics” |

| | | | | | |
|---|---|---|---|---|---|
| Descriptive data | 14 | (a) Give characteristics of study participants (*e.g.*, demographic, clinical, social) and information on exposures and potential confounders<br>(b) Indicate the number of participants with missing data for each variable of interest (c) *Cohort study*- summarise follow-up time (*e.g.*, average and total amount) | | | Described under "Patient characteristics"<br><br>Included in Supplementary B |
| Outcome data | 15 | *Cohort study*- Report numbers of outcome events or summary measures over time *Case-control study*- Report numbers in each exposure | | | |
| | | category, or summary measures of exposure *Cross-sectional study*- Report numbers of outcome events or summary measures | | | Numbers of cases listed in table 3 with summary demographics |

| | | | | | |
|---|---|---|---|---|---|
| Main results | 16 | (a) Give unadjusted estimates and, if applicable, confounder-adjusted estimates and their precision (e.g., 95% confidence interval). Make clear which confounders were adjusted for and why they were included (b) Report category boundaries when continuous variables were categorized<br>(c) If relevant, consider translating estimates of relative risk into absolute risk for a meaningful time period | | | Included under "Phenotypes"<br><br>Categorical boundaries for each cluster are reported in Figure 2 and the model results with pairwise comparisons included in Supplementary C |
| Other analyses | 17 | Report other analyses done— e.g., analyses of subgroups and interactions, and sensitivity analyses | | | No analyses of subgroups or interactions were included. Sensitivity analyses not performedDone |
| **Discussion** | | | | | |
| Key results | 18 | Summarise key results with reference to study objectives | | | First paragraph of Discussion |

| | | | | | |
|---|---|---|---|---|---|
| Limitations | 19 | Discuss limitations of the study, taking into account sources of potential bias or imprecision. Discuss both direction and magnitude of any potential bias | | RECORD 19.1: Discuss the implications of using data that were not created or collected to answer the specific research question(s). Include discussion of misclassification bias, unmeasured confounding, missing data, and changing eligibility over time, as they pertain to the study being reported. | Seventh paragraph of discussion and "Limitations" section within discussion |
| Interpretation | 20 | Give a cautious overall interpretation of results considering objectives, limitations, multiplicity of analyses, results from similar studies, and other relevant evidence | | | Second through seventh paragraph of discussion |
| Generalisability | 21 | Discuss the generalisability (external validity) of the study results | | | Discussed under "Limitations" |
| **Other Information** | | | | | |
| Funding | 22 | Give the source of funding and the role of the funders for the present study and, if applicable, for the original study on which the | | | Included under relevant heading |

| | | | | | |
|---|---|---|---|---|---|
| | | present article is based | | | |
| Accessibility of protocol, raw data, and programming code | | .. | | RECORD 22.1: Authors should provide information on how to access any supplemental information such as the study protocol, raw data, or programming code. | Supplemental information included with manuscript submission |

*Reference: Benchimol EI, Smeeth L, Guttmann A, Harron K, Moher D, Petersen I, Sørensen HT, von Elm E, Langan SM, the RECORD Working Committee. The REporting of studies Conducted using Observational Routinely-collected health Data (RECORD) Statement. *PLoS Medicine* 2015; in press.

*Checklist is protected under Creative Commons Attribution (CC BY) license.
