## Supplementary B for "Multidimensional analysis of preoperative patient-reported outcomes identifies distinct phenotypes in patients booked for total knee arthroplasty: Secondary analysis of the SHARKS registry in a metropolitan hospital department"

### Baseline analysis of knee osteoarthritis patients booked for total knee replacement

Corey Scholes

2024-01-19

**6 Export results** **27**

**References** **27**

#### 0.1 Preamble

**Author Affiliation:** EBM Analytics

**EBMAReference**: Pub_Arthroplasty_Baseline

**Version:** 1.0

### 1 Introduction

The following document outlines the preparation, assessment, cluster analysis and mode fitting conducted as a supplementary file for the paper (McGill et al. 2023).

### 2 Data Preparation

#### 2.1 Ready up libraries

#### 2.2 Import inputs

The project mastersheet was imported using the *openxlsx2* package (Barbone and Garbuszus 2024).

```
SHARKSSnapshot <- read_xlsx(
  file = "G:\\...QEII - registry sheet = "AllTimePoints",
  start_col = 3,
  col_names = TRUE,
  detect_dates = TRUE)
```

#### 2.3 Prepare dataset

The Mastersheet was prepared for the analysis using *tidyverse* (Wickham et al. 2019) to conduct the following actions;

- Drop columns
- Filter rows
- Rename columns

- Create new columns as required
  - Combine age columns
  - Calculate the period of time for each record input from the first record added in the series (in days)

```
## Create column list

Master <- SHARKSSnapshot %>% filter(
  InclusionSurgeryType == "Include" &
   !is.na(AgeAtExamination) &
   !is.na(ZipCode) &
   !is.na(Consent)
) %>%
  dplyr::select(
    PatientID,
    BilateralStatus,
    SurgeryDate,
    SurgeryRecordInputDate,
    AgeAtSurgery,
    AgeAtExamination,
    ZipCode,
    Gender,
    BMI,
    VR12MentalScore_Preop,
    VR12PhysicalScore_Preop,
    Oxford12_FunctionTotal_Preop,
    Oxford12_PainTotal_Preop
  ) %>%
  rename(PostCode = ZipCode,
         Sex = Gender,
         VR12MCS = VR12MentalScore_Preop,
         VR12PCS = VR12PhysicalScore_Preop,
         OKSFunction = Oxford12_FunctionTotal_Preop,
         OKSPain = Oxford12_PainTotal_Preop) %>%
  mutate(Age = ifelse(is.na(AgeAtExamination), AgeAtSurgery, AgeAtExamination),
         DaysFromInitial = as.numeric(difftime(SurgeryRecordInputDate, min(SurgeryRecordInpu
)
```

Socioeconomic status has been associated with preoperative (baseline) patient-reported outcomes in lower limb osteoarthritis patients receiving or due to receive arthroplasty surgery (Vega et al. 2022; Hoelen et al. 2023; Bonsel et al. 2023). The *haven* package (Wickham,

Miller, and Smith 2023) was used to import a previously prepared export from the Australian Tax Office , with the median total annual income (or loss) imported from the Stata file (.dta).

```
ATOFilePath <- "G:\\My Drive\\EBMA\\Client Drive\\QEII Jubilee\\EBMA Working\\Publications\\

# Read the Stata file into R
TaxableIncome <- read_dta(ATOFilePath)

Master1 <- left_join(Master,dplyr::select(TaxableIncome,mediantotalincomeorloss,postcode),
                     join_by(PostCode == postcode)) %>%
  mutate(MedianIncome = mediantotalincomeorloss/1000)
```

The adjusted Mastersheet was then used as input into the *gtsummary* package (Sjoberg et al. 2021) to create a table of patient characteristics and preoperative patient-reported outcomes.

```
Table1 <- tbl_summary(Master1, include = c(
  SurgeryRecordInputDate,
  DaysFromInitial,
  Age,
  Sex,
  BMI,
  BilateralStatus,
  MedianIncome,
  VR12MCS:OKSPain),
  missing_text = "Missing"
) %>%
  modify_header(
    label = "Characteristic",
    stat_0 = "Summary Statistics, N = 445"
  )

as_flex_table(Table1)
```

| Characteristic | Summary Statistics, N = 445[1] |
|---|---|
| SurgeryRecordInputDate | 2017-08-07 to 2023-10-09 |
| DaysFromInitial | 1,200 (763, 1,725) |
| Age | 69 (61, 75) |
| Sex | |
| Female | 234 (53%) |

| Characteristic | Summary Statistics, N = 445[1] |
|---|---|
| Male | 211 (47%) |
| BMI | 32.2 (28.3, 37.0) |
| Missing | 151 |
| BilateralStatus | |
| Bilateral | 156 (35%) |
| Unilateral | 289 (65%) |
| Median total income or loss | 48 (45, 51) |
| VR12MCS | 45 (37, 56) |
| Missing | 73 |
| VR12PCS | 28 (23, 34) |
| Missing | 73 |
| OKSFunction | 36 (29, 50) |
| Missing | 95 |
| OKSPain | 30 (20, 40) |
| Missing | 93 |

[1]Range; Median (IQR); n (%)

```
# print the column names that can be modified
##show_header_names(table1)
```

#### 3 Assess and address missingness

Missingness is an unavoidable characteristic of clinical datasets, in particular patients from non-english speaking or lower socioeconomic backgrounds are less likely to respond to patient-reported outcome measures in lower limb arthroplasty (Konopka et al. 2023). The Mastersheet was adjusted using *tidyverse to* include only the required columns and to convert categorical columns to factors. The *naniar* package (Tierney and Cook 2023) was used to generate a summary figure of missingness in the key variables of the analysis.

```
# Slice down Master1 slightly
Master2 <- Master1 %>% dplyr::select(!c(SurgeryDate,
```

```
                                    SurgeryRecordInputDate,
                                    AgeAtSurgery,
                                    AgeAtExamination,
                                    PostCode,
                                    mediantotalincomeorloss)) %>%
  mutate(
  BilateralStatus = as.factor(BilateralStatus),
  Sex = as.factor(Sex)
)

# Display the missing data pattern
Figure1a <- vis_miss(Master2)

Figure1b <- gg_miss_var(Master2,
            show_pct = TRUE)

Figure1a + Figure1b
```

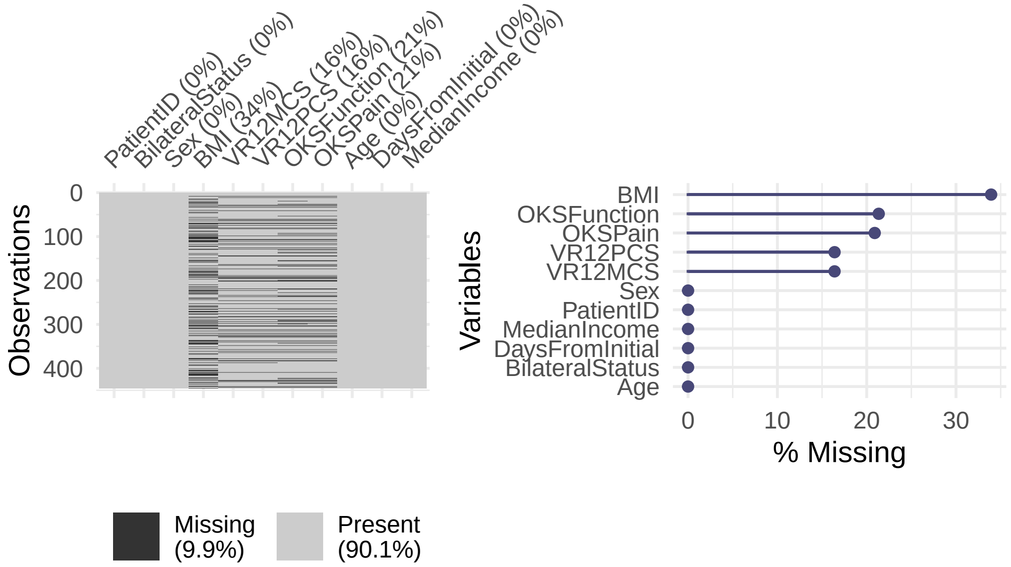

#### 3.1 Multiple Imputation 1

Multiple imputation using chained equations was implemented using the *mice* package (Buuren and Groothuis-Oudshoorn 2011). The predictor matrix was adjusted to exclude PatientID, a seed was set for reproducibility and 20 imputations selected.

```
Masterpred <- make.predictorMatrix(Master2)

# Switch off variables from predictormatrix
# Column
Masterpred[,"PatientID"] <- 0
# Row
Masterpred["PatientID",] <- 0

#Adjust MI sequence
Masterseq <- make.visitSequence(Master2)

# Perform multiple imputation with specified order and methods
Imputation1 <- mice::mice(
  data = Master2,
  m = 20,
  pred = Masterpred,
  seed = 4218,
  printFlag = FALSE
)

# Output imputed data in long format
Analysis_Data1 <- mice::complete(Imputation1,
                                 action = "long",
                                 include = TRUE)

#
```

A strip plot was generated to visualise model convergence for each variable.

```
plot(Imputation1,layout=c(2,5))
```

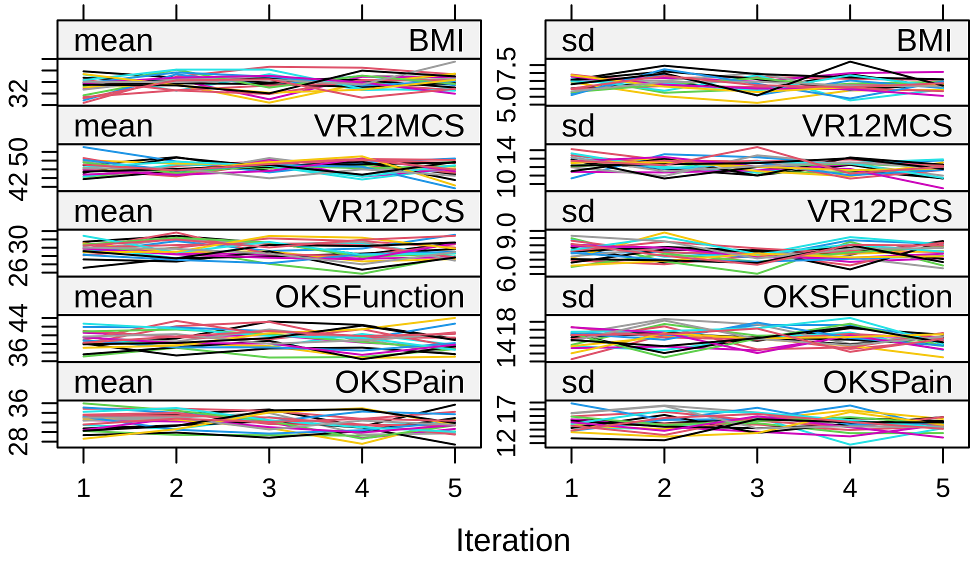

#### 4 Clustering Analysis

Conducting cluster analysis over multiple imputed data poses a significant challenge.

##### 4.1 Optimal cluster (k) selection

**Update 21-Jan-2024**: Found the *miclust* package (Basagaña et al. 2013), which identified 3 as the optimal cluster number, but it does not export any parameters for alternative cluster numbers other than the optimal selection based on *critcf* calculated within the package. For the purposes of this analysis I will continue with the more manual approach, which achieves the same outcome, but is not as elegant.

```
# Slice Down Dataset for clustering

Analysis_Data2a <- Analysis_Data1 %>% dplyr::select(.imp, VR12MCS, VR12PCS, OKSFunction, OKSP

#Convert to list
Analysis_List1 <- split(Analysis_Data2a, Analysis_Data2a$.imp)

# Remove .imp column from each data frame in the list
Analysis_List1 <- lapply(Analysis_List1, function(df) df[, !names(df) %in% ".imp"])
```

```
#Reformat data for clustering
Analysis_Data3 <- getdata(data = Analysis_List1)
```

```
#| label: Cluster Analysis1
#| include: false
#| echo: false
#| code-summary: "miclust cluster analysis"
#| warning: false
#|
ClusterK <- miclust(
  Analysis_Data3,
  method = "kmeans",
  search = "none",
  ks = 2:6,
  distance = "euclidean",
  centpos = "means",
  initcl = "rand",
  verbose = TRUE,
  seed = 4218
)
```

```
....imp 5....imp 10....imp 15....imp 20
 Analysis done.
```

```
plot(ClusterK, k = 4)
```

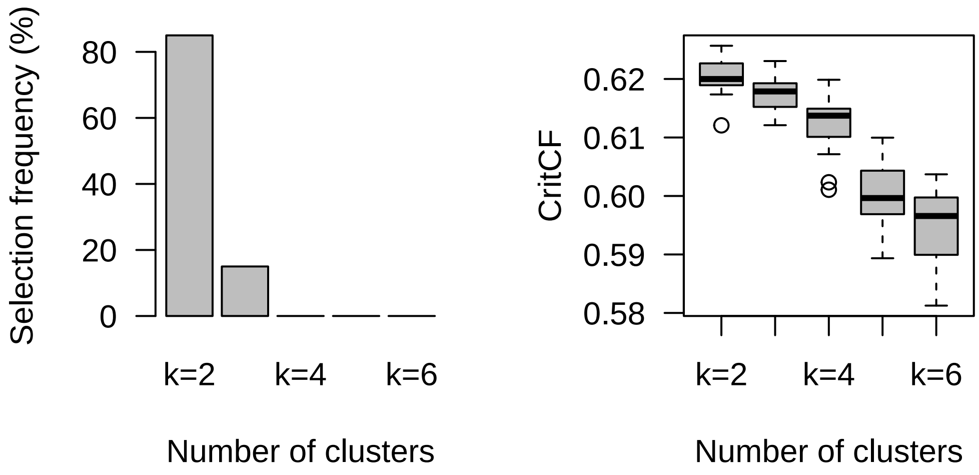

```
## summary forcing 3 clusters:
ClusterKsum <- summary(ClusterK, k = 3)
```

An attempt to identify an optimal cluster (phenotype) number using "elbow" method. Not able to achieve this over the imputed dataset, so had to reduce down to original dataset + m = 1. Used the *factoextra* package (Kassambara and Mundt 2020) to compare the total within-sum-squares across multiple options of k.

```
# optimal clusters
set.seed(4218)

# View imputed data (original with gaps filled by m = 1)
Master3a <- mice::complete(Imputation1)

fviz_nbclust(Master3a %>% dplyr::select(VR12MCS,
                                        VR12PCS,
                                        OKSFunction,
                                        OKSPain),
             kmeans,
             method = "wss") +
  geom_vline(xintercept = 3, linetype = 2)
```

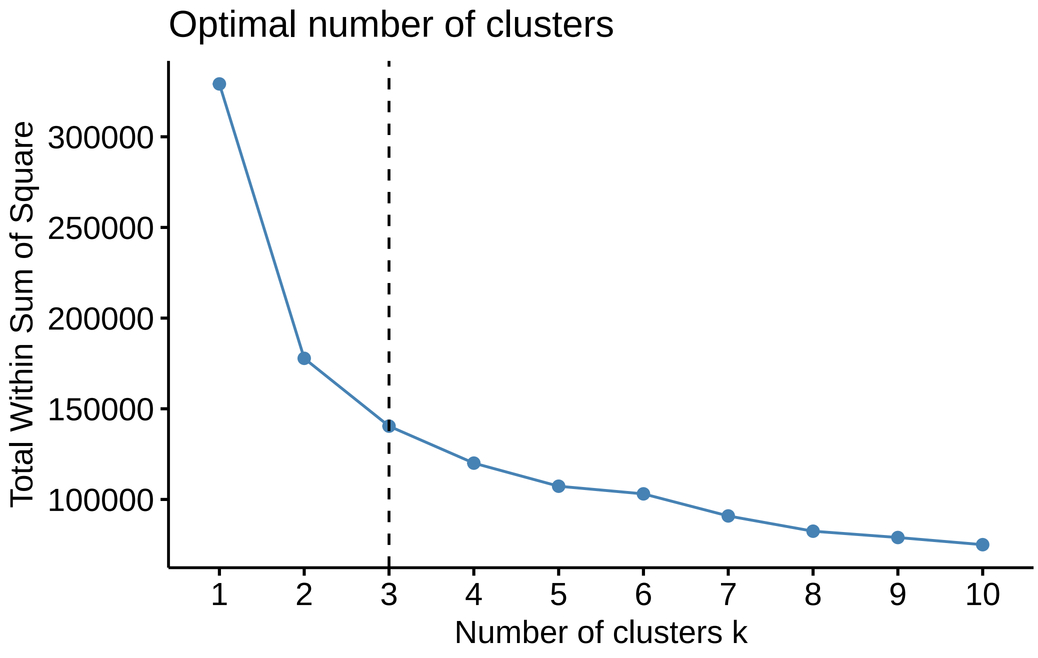

##### 4.2 Cluster Analysis

Still think there is merit in retaining 4 clusters (*phenotypes*) - within cluster sum squares is slightly reduced. Used base *R* to loop through each imputation subset of the imputed dataset and cluster based on the PROMs variables and then retrieving the cluster labels into a single dataframe.

```
# Initialize a dataframe to store cluster labels for each imputation
clusterlabels <- data.frame(matrix(NA, nrow = 445, ncol = 20))

#Convert Imputation1 to dataframe long without original data
Analysis_Data = mice::complete(Imputation1,
                               action = "long",
                               include = FALSE)

# Loop through each imputation
for (imp_index in 1:20) {

  set.seed(4218)

  # Extract the subset for the current imputation
  subset_data <- Analysis_Data %>%
```

```
    filter(.imp == imp_index)

  # Extract the variables for clustering
  cluster_variables <- subset_data %>%
    dplyr::select(VR12MCS, VR12PCS, OKSFunction, OKSPain)

  # Perform k-means clustering with k=4
  kmeans_result <- kmeans(cluster_variables, centers = 4)

  # Store the cluster assignments in the dataframe
  clusterlabels[, imp_index] <- kmeans_result$cluster
}

# Rename the columns for clarity
colnames(clusterlabels) <- paste(1:20, "Imp", sep = "_")
```

A heatmap was drawn to visualise the stability of cluster assignments over imputations.

```
heatmap(as.matrix(clusterlabels),
        Rowv = NA,
        Colv = NA,
        scale = "column",
        symm = TRUE,
        main = "heatmap Clusterlabels over Imputations",
        col = rainbow(4))

# Plot a corresponding legend
legend(x="right", legend=c("1", "2", "3","4"), fill = rainbow(4))
```

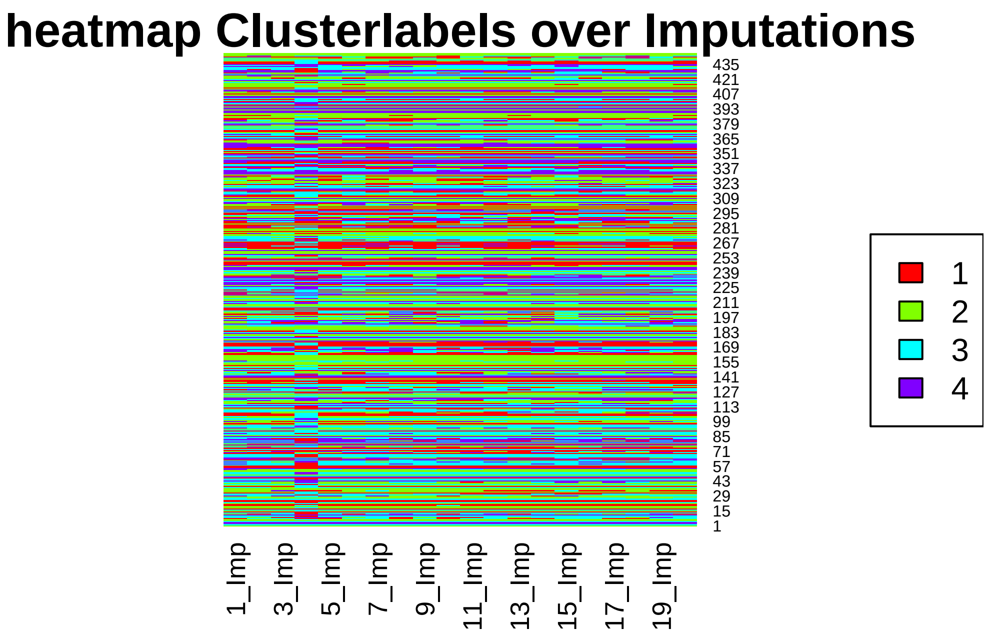

Used the *tidyverse* syntax to identify the modal value for each patient observation within the clusterlabels dataframe.

```
# Run from Clusterlabels
clusterlabels2 <- clusterlabels %>%
  rowwise() %>%
  mutate(ModeLabel = {
    tbl <- table(c_across(1:20))
    mode_values <- as.numeric(names(tbl)[tbl == max(tbl)])
    mode_values[1]
  })
```

The modal cluster labels (*phenotypes*) were added to the Mastersheet for further analysis. Based on an initial preview of the clustering and model fitting, the reference level for the cluster label (phenotype) was shifted to “2”.

```
Master3b <- Master3a %>% mutate(
  ModeLabel = as.factor(clusterlabels2$ModeLabel)
  ) %>%
      mutate(
    ModeLabel = relevel(ModeLabel, ref = "2")
  )
```

Taking into account the optimal selected k (3), the results of both 3-cluster and 4-cluster results were compared using a plotting function from *factoextra* package that performed a principal component analysis on the cluster results, before plotting the centred data.

```
kmeans_short3 <- kmeans(Master3b %>% dplyr::select(VR12MCS,VR12PCS,OKSFunction,OKSPain),
                       centers = 3,
                       iter.max = 10,
                       nstart = 6
                       )

kmeans_short4 <- kmeans(Master3b %>% dplyr::select(VR12MCS,VR12PCS,OKSFunction,OKSPain),
                       centers = 4,
                       iter.max = 10,
                       nstart = 8
                       )

ClusterPlot1 <- fviz_cluster(
  kmeans_short3,
  data = Master3b %>% dplyr::select(VR12MCS,VR12PCS,OKSFunction,OKSPain)
)

ClusterPlot2 <- fviz_cluster(
  kmeans_short4,
  data = Master3b %>% dplyr::select(VR12MCS,VR12PCS,OKSFunction,OKSPain)
)

ClusterPlot1 + ClusterPlot2
```

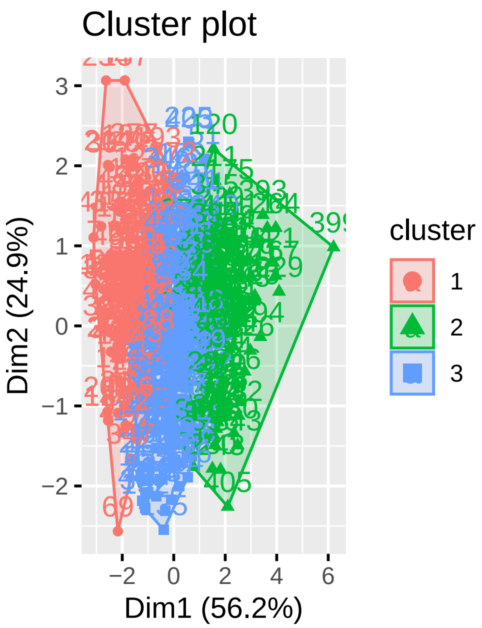

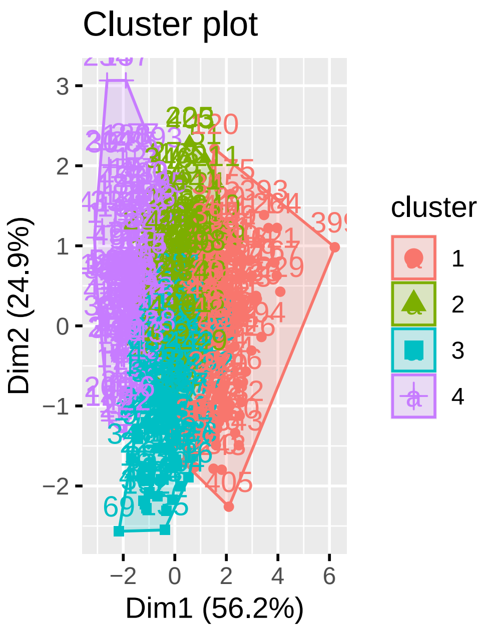

A second plot was created to recreate using the modal label from imputed analysis

```
# Select the columns of interest
selected_columns <- c("VR12MCS", "VR12PCS", "OKSFunction", "OKSPain", "ModeLabel")
Master4 <- Master3b[, selected_columns]

# Perform PCA
pca_result <- prcomp(Master4[, -which(names(Master4) == "ModeLabel")], scale. = TRUE)

# Combine PCA results with cluster labels
pca_with_clusters <- cbind(as.data.frame(predict(pca_result)), ModeLabel = Master4$ModeLabel)

# Create a ggplot using ggfortify
ggplot(pca_with_clusters, aes(x = PC1, y = PC2, color = as.factor(ModeLabel))) +
  geom_point() +
  labs(title = "PCA Visualization with Cluster Labels",
       x = "Principal Component 1",
       y = "Principal Component 2") +
  theme_minimal() +
  scale_color_ordinal()
```

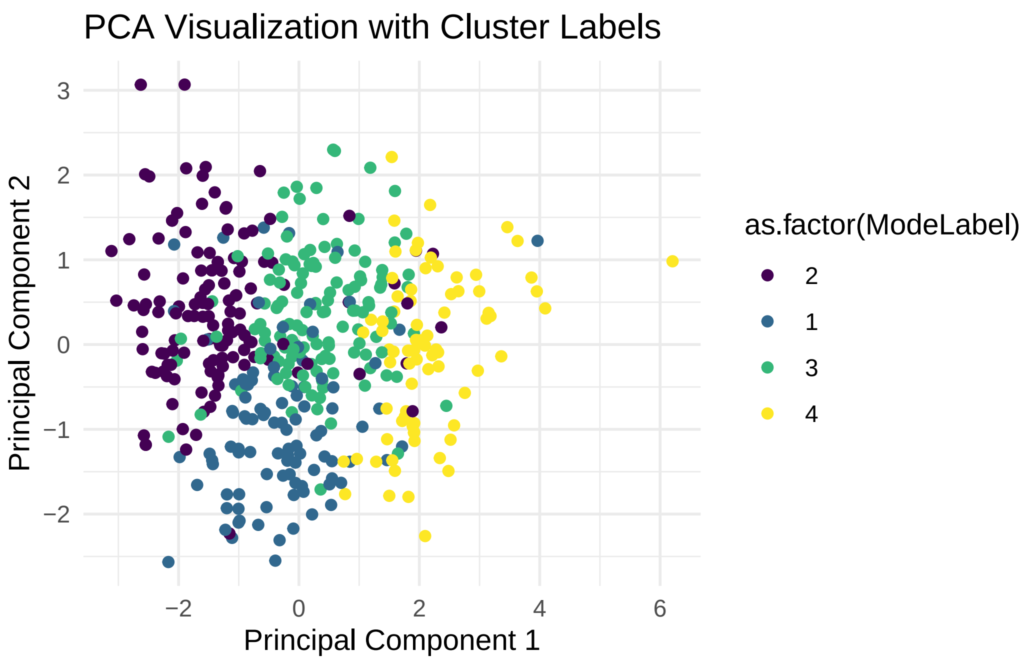

The results were also visualised using the *scatterplot3d* package (Ligges and Machler 2003) to illustrate the separation between clusters (phenotypes) with a 3-dimensional view.

```
plot_3d <- scatterplot3d(x = Master4$VR12MCS,
                         y = Master4$OKSFunction,
                         z = Master4$OKSPain,
                         col.axis = "black",
                         col.grid = "cyan",
                         pch = 20,
                         #highlight.3d = TRUE
                         color = Master4$ModeLabel,
                         angle = 50,
                         xlab = "VR12MCS",
                         ylab = "OKSFunction",
                         zlab = "OKSPain"
                         )
```

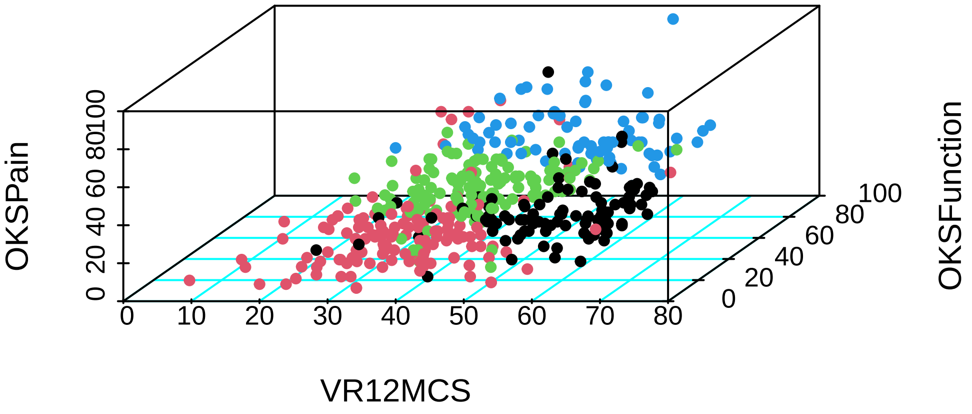

It is now possible to turn attention to model-fitting on the imputed dataset.

### 5 Model fitting

#### 5.1 Multiple imputation 2

To facilitate model fitting after imputation, it is important that the dependent variable is included in the imputation model (Ginkel et al. 2019), so that now the phenotype labels have been after the initial imputation, a second round is necessary to reduce bias inherent in performing model fitting with the dependent variable that was not included as part of the mi input.

```
Masterpred2 <- make.predictorMatrix(Master3b)

# Switch off variables from predictormatrix
# Column
Masterpred2[,"PatientID"] <- 0
# Row
Masterpred2["PatientID",] <- 0

# Column
```

```
#Masterpred2[,"ModeLabel"] <- 0
# Row
#Masterpred2["ModeLabel",] <- 0
```

```
# rerun mice

#Add modelabel to original dataset

Master5 <- Master2 %>% mutate(ModeLabel = Master3b$ModeLabel)

#Adjust MI sequence
Masterseq2 <- make.visitSequence(Master5)

# Perform multiple imputation with specified order and methods
Imputation2 <- mice::mice(
  data = Master5,
  m = 20,
  pred = Masterpred2,
  seed = 4218,
  printFlag = FALSE
)

# View imputed data (long)
Imputation2_Data <- mice::complete(Imputation2, "long")
# Imputed data (short)
Master6 <- mice::complete(Imputation2)
```

##### 5.2 Patient-reported outcomes

To illustrate for the reader the separation between clusters for the input variables, model-fitting was performed using the imputed datasets. The *Imputation2_Data* dataframe was used as an input into the models.

```
# Run linear models

modelVR12MCSimp = with(Imputation2, exp = lm(VR12MCS ~ factor(ModeLabel)))
modelVR12PCSimp = with(Imputation2, exp = lm(VR12PCS ~ factor(ModeLabel)))
modelOKSFunctionimp = with(Imputation2, exp = lm(OKSFunction ~ factor(ModeLabel)))
modelOKSPainimp = with(Imputation2, exp = lm(OKSPain ~ factor(ModeLabel)))
```

The *gtsummary* package was used to create summary tables for each model and combine them into one cohesive table for presentation.

```
#| label: linear regression
#| include: false
#| echo: false
#| code-summary: "Linear regression summaries"
#| warning: false

Table2Vr12MCS <- tbl_regression(modelVR12MCSimp,tidy_fun = pool_and_tidy_mice)
```

```
pool_and_tidy_mice(): Tidying mice model with
`mice::pool(x) %>% mice::tidy(conf.int = TRUE, conf.level = 0.95, exponentiate = FALSE)`
```

```
Table2Vr12PCS <- tbl_regression(modelVR12PCSimp,tidy_fun = pool_and_tidy_mice)
```

```
pool_and_tidy_mice(): Tidying mice model with
`mice::pool(x) %>% mice::tidy(conf.int = TRUE, conf.level = 0.95, exponentiate = FALSE)`
```

```
Table2OKSFunction <- tbl_regression(modelOKSFunctionimp,tidy_fun = pool_and_tidy_mice)
```

```
pool_and_tidy_mice(): Tidying mice model with
`mice::pool(x) %>% mice::tidy(conf.int = TRUE, conf.level = 0.95, exponentiate = FALSE)`
```

```
Table2OKSPain <- tbl_regression(modelOKSPainimp,tidy_fun = pool_and_tidy_mice)
```

```
pool_and_tidy_mice(): Tidying mice model with
`mice::pool(x) %>% mice::tidy(conf.int = TRUE, conf.level = 0.95, exponentiate = FALSE)`
```

```
Table2 <- tbl_merge(tbls = list(Table2Vr12MCS,Table2Vr12PCS,Table2OKSFunction,Table2OKSPain)
                              tab_spanner = c("VR12MCS", "VR12PCS","OKSFunction","OKSPain"))

knitr::knit_print(Table2)
```

```
Table printed with `knitr::kable()`, not {gt}. Learn why at
https://www.danieldsjoberg.com/gtsummary/articles/rmarkdown.html
To suppress this message, include `message = FALSE` in code chunk header.
```

| Characteristic | Beta | 95% CI | p-value | Beta | 95% CI | p-value | Beta | 95% CI | p-value | Beta | 95% CI | p-value |
|---|---|---|---|---|---|---|---|---|---|---|---|---|
| factor(ModeLabel) | | | | | | | | | | | | |
| 2 | — | — | | — | — | | — | — | | — | — | |
| 1 | 25 | 22, 27 | <0.001 | -0.31 | -2.1, 1.5 | 0.7 | 11 | 8.0, 14 | <0.001 | 10 | 7.5, 13 | <0.001 |
| 3 | 8.0 | 5.7, 10 | <0.001 | 5.9 | 4.2, 7.6 | <0.001 | 21 | 18, 23 | <0.001 | 19 | 17, 21 | <0.001 |
| 4 | 19 | 17, 22 | <0.001 | 11 | 8.9, 13 | <0.001 | 40 | 37, 42 | <0.001 | 37 | 35, 40 | <0.001 |

A "predict then combine" approach was implemented based on the recommendations of Miles (2015), using code presented in (Solomon 2021). The code was converted to *tidyverse* syntax for clarity. The *purrr* package (Wickham and Henry 2023) was used to map across dataframes within a list variable.

```
#Setup a constant
m <- 20

fitted_linesVR12MCS <-
  tibble(.imp = 1:20) %>%
  mutate(p = map(.imp, ~ predict.lm(modelVR12MCSimp$analyses[[.]], se.fit = TRUE) %>%
  data.frame())) %>%
  unnest(p) %>%
  dplyr::select(!c(df, residual.scale)) %>%
  mutate(
    modelabel = Imputation2_Data$ModeLabel
  ) %>%
  group_by(modelabel) %>%
  summarise(fit_bar = mean(fit),
            v_w     = mean(se.fit^2),
            v_b     = sum((fit - fit_bar)^2) / (m - 1),
            v_p     = v_w + v_b * (1 + (1 / m)),
            se_p    = sqrt(v_p)) %>%
  # use the _p suffix to indicate these are pooled
  mutate(lwr_p = fit_bar - se_p * 1.96,
         upr_p = fit_bar + se_p * 1.96)

fitted_linesVR12PCS <-
```

```
  tibble(.imp = 1:20) %>%
  mutate(p = map(.imp, ~ predict.lm(modelVR12PCSimp$analyses[[.]], se.fit = TRUE) %>%
  data.frame())) %>%
  unnest(p) %>%
  dplyr::select(!c(df, residual.scale)) %>%
  mutate(
    modelabel = Imputation2_Data$ModeLabel
  ) %>%
  group_by(modelabel) %>%
  summarise(fit_bar = mean(fit),
            v_w     = mean(se.fit^2),
            v_b     = sum((fit - fit_bar)^2) / (m - 1),
            v_p     = v_w + v_b * (1 + (1 / m)),
            se_p    = sqrt(v_p)) %>%
  # use the _p suffix to indicate these are pooled
  mutate(lwr_p = fit_bar - se_p * 1.96,
         upr_p = fit_bar + se_p * 1.96)

fitted_linesOKSFunction <-
  tibble(.imp = 1:20) %>%
  mutate(p = map(.imp, ~ predict.lm(modelOKSFunctionimp$analyses[[.]], se.fit = TRUE) %>%
  data.frame())) %>%
  unnest(p) %>%
  dplyr::select(!c(df, residual.scale)) %>%
  mutate(
    modelabel = Imputation2_Data$ModeLabel
  ) %>%
  group_by(modelabel) %>%
  summarise(fit_bar = mean(fit),
            v_w     = mean(se.fit^2),
            v_b     = sum((fit - fit_bar)^2) / (m - 1),
            v_p     = v_w + v_b * (1 + (1 / m)),
            se_p    = sqrt(v_p)) %>%
  # use the _p suffix to indicate these are pooled
  mutate(lwr_p = fit_bar - se_p * 1.96,
         upr_p = fit_bar + se_p * 1.96)

fitted_linesOKSPain <-
  tibble(.imp = 1:20) %>%
  mutate(p = map(.imp, ~ predict.lm(modelOKSPainimp$analyses[[.]], se.fit = TRUE) %>%
  data.frame())) %>%
  unnest(p) %>%
```

```r
  dplyr::select(!c(df, residual.scale)) %>%
  mutate(
    modelabel = Imputation2_Data$ModeLabel
  ) %>%
  group_by(modelabel) %>%
  summarise(fit_bar = mean(fit),
            v_w     = mean(se.fit^2),
            v_b     = sum((fit - fit_bar)^2) / (m - 1),
            v_p     = v_w + v_b * (1 + (1 / m)),
            se_p    = sqrt(v_p)) %>%
  # use the _p suffix to indicate these are pooled
  mutate(lwr_p = fit_bar - se_p * 1.96,
         upr_p = fit_bar + se_p * 1.96)
```

Shifted the reference phenotype (2) to the left for clarity. The *ggplot2* package (Wickham 2016) was used to generate box plots for each PROM input into the cluster model and then combined into one figure using *patchwork* (Pedersen 2023).

```r
#Shifted reference phe

# Assuming you have a dataframe named fitted_lines2

# Create a boxplot with upper and lower confidence intervals
plotVR12MCS5imp <- ggplot(fitted_linesVR12MCS, aes(x = modelabel, y = fit_bar)) +
  geom_boxplot(width = 0.5, fill = "lightblue", color = "black", alpha = 0.7) +
  geom_errorbar(aes(ymin = lwr_p, ymax = upr_p), width = 0.2, position = position_dodge(0.5)
  labs(x = "Phenotype",
       y = "VR12MCS Mean") +
  theme_minimal()

plotVR12PCS5imp <- ggplot(fitted_linesVR12PCS, aes(x = modelabel, y = fit_bar)) +
  geom_boxplot(width = 0.5, fill = "lightblue", color = "black", alpha = 0.7) +
  geom_errorbar(aes(ymin = lwr_p, ymax = upr_p), width = 0.2, position = position_dodge(0.5)
  labs(x = "Phenotype",
       y = "VR12PCS Mean") +
  theme_minimal()

plotOKSFunction5imp <- ggplot(fitted_linesOKSFunction, aes(x = modelabel, y = fit_bar)) +
  geom_boxplot(width = 0.5, fill = "lightblue", color = "black", alpha = 0.7) +
  geom_errorbar(aes(ymin = lwr_p, ymax = upr_p), width = 0.2, position = position_dodge(0.5)
  labs(x = "Phenotype",
       y = "OKSFunction Mean") +
```

```
  theme_minimal()

plotOKSPain5imp <- ggplot(fitted_linesOKSPain, aes(x = modelabel, y = fit_bar)) +
  geom_boxplot(width = 0.5, fill = "lightblue", color = "black", alpha = 0.7) +
  geom_errorbar(aes(ymin = lwr_p, ymax = upr_p), width = 0.2, position = position_dodge(0.5)
  labs(x = "Phenotype",
       y = "OKSPain Mean") +
  theme_minimal()

plotVR12MCS5imp + plotVR12PCS5imp + plotOKSFunction5imp + plotOKSPain5imp +
  plot_annotation(
  title = "Pooled predicted estimates of PROMs",
  subtitle = "VR12MCS, VR12PCS, OKSFunction, OKSPain")
```

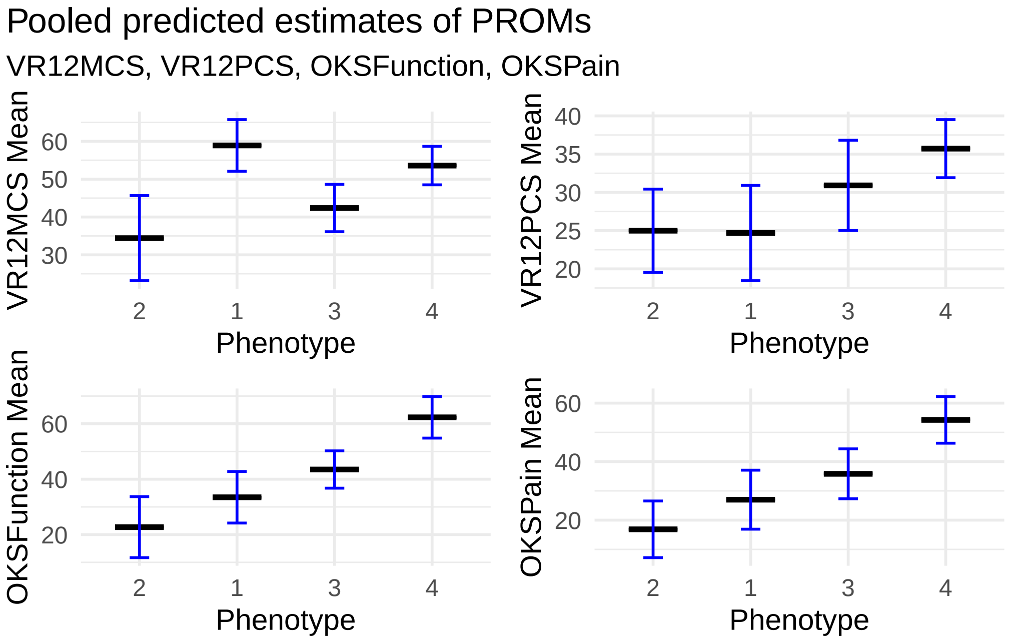

#### 5.3 Phenotype and patient characteristics

Due to the lack of assumptions regarding the total ordering of the *phenotypes* (other than 2 being the reference category for comparisons) a multinomial logistic regression model was chosen to assess the relationship between patient characteristics and *phenotype.* The *nnet* package (Venables and Ripley 2002) was selected as the most compatible with the imputed dataset.

The pooled model estimates were extracted using the *gtsummary* package.

```
#| label: logistic regression summary
#| include: false
#| echo: false
#| code-summary: "Multinomial logistic regression summary"
#| warning: false

# Summarize the model
Table3 <- tbl_regression(multinom_cluster4,
                                add_estimate_to_reference_rows = TRUE,
                                exponentiate = TRUE,
                                show_single_row = c("Sex","BilateralStatus"),
                                label = list(Sex ~ "Male",
                                             BilateralStatus ~ "Bilateral",
                                             BMI ~ "Body Mass Index",
                                             MedianIncome ~ "MedianIncome"
                                              ),
                              tidy_fun = pool_and_tidy_mice
                             )
```

```
pool_and_tidy_mice(): Tidying mice model with
`mice::pool(x) %>% mice::tidy(conf.int = TRUE, conf.level = 0.95, exponentiate = TRUE)`
i Multinomial models have a different underlying structure than the models
gtsummary was designed for. Other gtsummary functions designed to work with
tbl_regression objects may yield unexpected results.
```

```
knitr::knit_print(Table3)
```

```
Table printed with `knitr::kable()`, not {gt}. Learn why at
https://www.danieldsjoberg.com/gtsummary/articles/rmarkdown.html
To suppress this message, include `message = FALSE` in code chunk header.
```

| **Outcome** | **Characteristic** | **OR** | **95% CI** | **p-value** |
|---|---|---|---|---|
| 1 | Age | 1.08 | 1.04, 1.12 | <0.001 |
| | Male | 1.98 | 1.13, 3.49 | 0.018 |
| | Body Mass Index | 0.96 | 0.91, 1.02 | 0.2 |
| | Bilateral | 1.16 | 0.66, 2.04 | 0.6 |
| | MedianIncome | 0.98 | 0.94, 1.03 | 0.4 |

| Outcome | Characteristic | OR | 95% CI | p-value |
|---|---|---|---|---|
| 3 | Age | 1.04 | 1.01, 1.07 | 0.019 |
| | Male | 1.51 | 0.89, 2.56 | 0.13 |
| | Body Mass Index | 0.93 | 0.88, 0.98 | 0.011 |
| | Bilateral | 2.16 | 1.24, 3.76 | 0.006 |
| | MedianIncome | 1.07 | 1.02, 1.12 | 0.006 |
| 4 | Age | 1.06 | 1.03, 1.10 | <0.001 |
| | Male | 1.95 | 1.06, 3.58 | 0.031 |
| | Body Mass Index | 0.94 | 0.88, 1.00 | 0.047 |
| | Bilateral | 1.57 | 0.84, 2.92 | 0.2 |
| | MedianIncome | 1.05 | 1.00, 1.11 | 0.052 |

The conversion between model estimates and summary statistics across the imputations proved to be quite problematic. To my knowledge, there is no reliable method readily available to produce fits, standard error of fits and confidence intervals for multinomial regression models, as there are (*predict*) for linear fits of continuous outcomes. While it is possible to extract the fitted values from the multinomial logistic regression results for each imputation, the errors must be calculated. For the sake of time, a shortcut was implemented that generated the summary statistics from the imputed data (original + m =1).

```
# Summarise relationship between predictors and ModeLabel

Table4 <- tbl_summary(Master6, include = c(
  Age,
  Sex,
  BMI,
  BilateralStatus,
  MedianIncome,
  ModeLabel),
  by = ModeLabel,
  type = list(
    BilateralStatus ~ "dichotomous",
    Sex ~ "dichotomous"),
  value = list(
    BilateralStatus ~ "Bilateral",
    Sex ~ "Female"),
  label = list(
    Age ~ "Age (Years)",
    Sex ~ "Female",
    BilateralStatus ~ "Bilateral",
```

```r
    BMI ~ "Body Mass Index (kgm^2)",
    MedianIncome ~ "MedianIncome (1000s/pa)"
    ),
  statistic = list(all_continuous() ~ "{mean} ({sd})",
                   all_categorical() ~ "{p}"
    )
  ) %>%
  add_ci(statistic = list(all_continuous() ~ "{conf.low} - {conf.high}",
                          all_categorical() ~ "{conf.low} - {conf.high}")
  ) %>%
  add_stat_label(location = "row") %>%
  modify_header(
  label = "Characteristic",
  stat_1 = "Phenotype 2 (REF), N = 134",
  ci_stat_1 = "95%CI",
  stat_2 = "Phenotype 1, N = 101",
  ci_stat_2 = "95%CI",
  stat_3 = "Phenotype 3, N = 132",
  ci_stat_3 = "95%CI",
  stat_4 = "Phenotype 4, N = 78",
  ci_stat_4 = "95%CI"
  ) %>%
    modify_footnote(all_stat_cols() ~ "REF = Reference Category")

as_flex_table(Table4)
```

| Characteristic | Phenotype 2 (REF), N = 134[1] | 95%CI[2] | Phenotype 1, N = 10 |
|---|---|---|---|
| Age (Years), Mean (SD) | 64 (10) | 63 - 66 | 71 (8) |
| Female, % | 63 | 54 - 71 | 48 |
| Body Mass Index (kgm ^2), Mean (SD) | 36 (7) | 35 - 37 | 32 (6) |
| Bilateral, % | 43 | 35 - 52 | 39 |
| MedianIncome (1000s/pa), Mean (SD) | 46 (5) | 45 - 47 | 46 (6) |

[1]REF = Reference Category

[2]CI = Confidence Interval

#### 6 Export results

The Table results were exported to png format using the *gt* package (Iannone et al. 2023).

```
Table1 %>% as_gt() %>% gtsave(filename = "Table1.png")
Table2 %>% as_gt() %>% gtsave(filename = "Table2.png")
Table3 %>% as_gt() %>% gtsave(filename = "Table3.png")
Table4 %>% as_gt() %>% gtsave(filename = "Table4.png")
```
